# Effectiveness of Social Networks in Enhancing the Uptake of Artemisinin-based Combination Therapy (ACT) in Malaria-Prone Zones of Kenya

**DOI:** 10.1101/2025.02.04.25321665

**Authors:** Florence Nelima Nyongesa, John Kamau Gathiaka, Germano Mwabu

## Abstract

**Introduction:** Malaria remains a critical health challenge in Kenya, particularly among children under five years who are highly vulnerable to its complications. Artemisinin-based combination therapies (ACTs) are the recommended first-line treatment for uncomplicated malaria. However, their uptake varies due to socio-economic inequalities, geographic disparities, and the influence of social networks. Understanding how social interactions, particularly those mediated by religious affiliations, impact ACT uptake can inform strategies to improve malaria treatment outcomes in malaria-prone regions.

**Methods:** This study utilized data from the Kenya Malaria Indicator Survey (KMIS) 2020, employing a cross-sectional design and stratified two-stage cluster sampling. Data on malaria prevalence, treatment-seeking behavior, and ACT usage were analyzed using descriptive statistics and logistic regression. The analysis focused on socio-demographic factors, geographic differences, and the role of social networks mediated through religious affiliations.

**Results:** ACT uptake among children under five was 52%. Male children were less likely to receive ACTs, with probabilities 6.3% lower in rural areas and 3.1% lower in urban areas. Younger children in rural areas, particularly those aged one, had a 5.7% higher likelihood of ACT usage, while uptake declined with age. Caregivers’ education significantly enhanced ACT uptake in rural areas, increasing the likelihood by 25%. Rural Muslim households were 17.2% more likely to use ACTs, while urban Christian households showed modest improvements. Wealth disparities also affected uptake, with wealthier urban households less likely to use ACTs.

**Conclusion:** The study highlights the critical role of social networks, particularly religious affiliations, in shaping ACT uptake in malaria-prone zones of Kenya. Addressing barriers to access through these networks offers a promising avenue for increasing ACT utilization and improving health outcomes. Leveraging community-driven approaches and religious institutions could enhance equitable malaria treatment coverage across vulnerable populations.

### What is already known on this topic

Caregiver education has been consistently identified as a key factor in improving health service utilization, including malaria treatment, due to increased awareness and understanding of the benefits of ACT.

### What this study adds

The study offers new evidence on how social networks, particularly religious affiliations (Christian and Muslim), influence the uptake of ACTs among children under five in malaria-prone areas. This highlights the potential of leveraging community and religious structures for health interventions.

### How this study might affect research, practice or policy

This study advances research by emphasizing the critical role of social networks, particularly religious affiliations, in shaping health behaviors, providing a basis for future interdisciplinary investigations into the dynamics of community influence on malaria treatment. It informs practice by highlighting the potential of engaging religious and community leaders to promote ACT uptake and designing tailored interventions addressing disparities in education and wealth. For policy, the findings advocate for leveraging religious partnerships to amplify health messaging, prioritizing targeted resources to reduce socio-economic and geographic barriers, and integrating community-driven approaches into malaria control strategies to achieve equitable health outcomes.

## Background

Malaria remains a critical public health issue in sub-Saharan Africa, bearing the brunt of global malaria cases and deaths. In Kenya, it is a leading cause of illness, particularly in the western and coastal regions, despite advancements in treatment availability. Artemisinin-based Combination Therapies (ACTs) are the first-line treatment for uncomplicated malaria, but their effectiveness depends significantly on community-level uptake and appropriate use. [1] Research by Hetzel et al. (2008) [2] highlights barriers to effective malaria treatment in rural Tanzania, such as limited healthcare access, financial constraints, inadequate knowledge, and antimalarial drug stockouts. These challenges persist despite community health worker initiatives, emphasizing the need for integrated approaches addressing systemic healthcare gaps and enhancing community-level education to boost treatment uptake and reduce malaria-related morbidity and mortality.

Social interactions within family networks, peer groups, and local communities significantly influence health behaviors, including ACT uptake. Studies demonstrate the importance of social networks in shaping treatment-seeking behaviors, with peer influence, community norms, and family advice playing vital roles in malaria treatment decisions. [3,4] In Kenya, social dynamics, particularly through religious, social, and community networks, strongly impact malaria treatment demand. [5] Individuals involved in social groups, such as women’s or church-based organizations, are more likely to seek timely and effective treatment. Additionally, community health workers embedded within these networks serve as crucial links between healthcare systems and local communities, disseminating accurate health information to improve ACT utilization.

However, challenges persist, including misinformation spread through social networks, which can lead to reliance on ineffective treatments or delayed ACT use. [6] Addressing these issues requires strategies that amplify positive social influences while mitigating harmful practices. This study focuses on understanding the role of social interactions, particularly religious and community affiliations, in influencing ACT demand and utilization in Kenya. [7] By examining how group-level dynamics affect treatment-seeking behavior, it aims to uncover factors that either facilitate or hinder ACT uptake.

Understanding the interplay of community dynamics in shaping individual health behaviors is vital for designing culturally relevant and accessible malaria interventions. By leveraging social networks, this research seeks to enhance the effectiveness of malaria control programs, particularly among vulnerable groups like children under five. The findings aim to inform policy and practice, ensuring that treatment strategies are not only effective but also contextually appropriate to the needs of at-risk populations. [8]

### Literature

The empirical literature on the usage of ACTs among children under five in malaria-endemic regions offers a comprehensive view of the multifaceted factors influencing treatment decisions. Research by Chanda et al. (2011) ^[9]^ underscores the critical role of supply chain management, including drug availability and accessibility, in determining ACT utilization in Zambia. Likewise, Cissé et al. (2016) ^[10]^ emphasize the role of socio-demographic factors, caregiver knowledge, and access to healthcare in rural Uganda and Senegal, respectively, underscoring how these factors greatly influence patterns of ACT utilization. This study also highlights the importance of community-based education and targeted interventions that tackle both socio-economic and educational challenges. These findings suggest that improving caregiver awareness and attitudes toward malaria treatment, as well as ensuring adherence to prescribed medication regimens, are crucial for enhancing ACT uptake and effectiveness. ^[10]^

Moreover, the persistence of traditional medicine use, as explored by Namukobe et al. (2011) and Okebe et al. (2014) ^[11,12]^, reveals a complex landscape where ACTs are often used alongside or replaced by alternative treatments. This highlights the need for culturally sensitive health education programs that promote the appropriate use of ACTs while respecting local traditions.

Mitiku and Assefa (2017) and Ajumobi et al., (2017) ^[13,14]^ studies examined caregivers’ perception of malaria and treatment-seeking behaviour for under five children. The research explores the use of various medicines, including traditional remedies and over-the-counter drugs, in households with children under five. The findings reveal a complex landscape of treatment practices, where ACTs are sometimes supplemented or replaced by alternative medicines due to factors such as cost, accessibility, and cultural beliefs.

In general, these empirical studies emphasize the need for a comprehensive approach to malaria treatment that tackles supply chain challenges, improves caregiver education, and combines traditional practices with modern medical treatments. This comprehensive understanding is crucial for formulating public health strategies that enhance malaria treatment effectiveness and reduce the burden of the disease among vulnerable populations in malaria-endemic regions.

## Methodology

### Theoretical Framework

The decision-making process of child caregivers regarding malaria treatment can be modeled by integrating the health demand framework developed by Grossman (1972) ^[15]^ with the economic principles outlined by Phelps (2016). ^[16]^ This integrated model provides a comprehensive understanding of how caregivers allocate resources and make treatment choices in the context of childhood malaria.

Grossman’s model conceptualizes health as a type of durable capital that produces a flow of healthy time, from which individuals derive benefit or utility. In the context of child caregivers, the health of their child can be seen as part of this capital stock. Caregivers must decide how much to invest in maintaining or improving this stock through healthcare actions, such as seeking malaria treatment.

The key components of this model are the Health Capital, (H) which is the current level of the child’s health, which can be increased or maintained through healthcare investments (e.g., purchasing and administering ACTs); the Utility Function, (U), where the caregivers derive utility from the health of their child, which is influenced by the child’s health status and other factors such as financial resources and time; and caregiver’s allocation of resources (C), including time, money, and effort, to maintain or improve their child’s health. This includes decisions related to seeking and administering malaria treatment.

In Grossman’s model, the utility function U depends on two primary factors:

Health Capital (H): The child’s health, which contributes to the caregiver’s overall utility. Consumption of Other Goods (C): Other goods or services that the caregiver consumes, which also contribute to utility.

The utility function can be represented as:

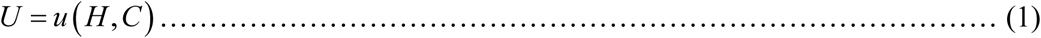

Where:

H denotes the health of the child, influenced by the caregiver’s investment in healthcare (e.g., ACTs).

C represents the consumption of all other goods, constrained by the caregiver’s income and the cost of healthcare.

Following the assumptions of Rosenzweig and Shultz (1983) and Mwabu (2008), we posit that the health production function for children is integrated into the utility-maximizing behavior of the caregiver. Health (H) is generated through investments in healthcare, such as the use of malaria treatment commodities like ACTs.

Health production function can be represented as:

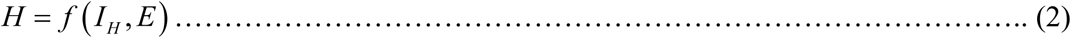

Where:

I_H_ represents the caregiver’s investment in healthcare (e.g., purchasing ACTs, time spent on treatment).

E represents external factors such as environmental conditions, baseline health, and genetic predispositions.

The child caregivers maximize equations (1) and (2) facing a budget constraint in equation (3) where their income Y is allocated between spending on healthcare I_H_ and other goods C:

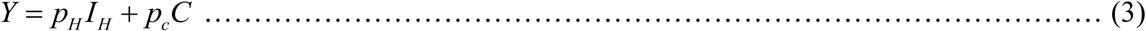

Where:

pH represents the cost of healthcare investments, such as the price of ACTs. p_C_ refers to the cost of other goods.

This budget constraint can be rewritten as:

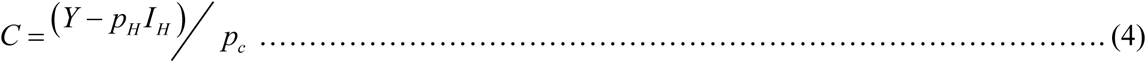

Caregivers aim to maximize their utility by choosing the optimal level of healthcare investment I_H_, given their budget constraints.

The utility optimization problem can be formulated as:

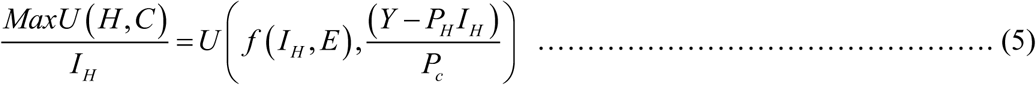

To incorporate social interactions, we add a parameter S representing the influence of social factors (peer influence, community norms, information exchange):

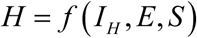

The final utility function, incorporating health capital, consumption, and social interactions, is:

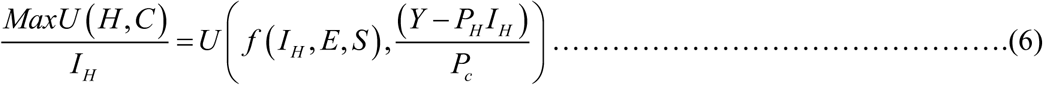

This function illustrates that the caregiver’s utility depends on the health of their child (influenced by investments in malaria treatment and social factors) and their ability to consume other goods within their budget. The caregiver will choose the level of investment I_H_ that maximizes their utility, balancing the costs of healthcare with the benefits of improved health and the consumption of other goods.

### Analytical Model

This study aims to assess the impact of social interactions on the demand and use of treatment commodities in malaria-endemic regions of Kenya. Since the dependent variable is discrete, we employ the binary logit regression model in examining the social interaction effects on demand and utilization of malaria treatment commodities.

First, we estimate the binary logit regression model in equation (7) below for the demand of ACT/other malaria medicines without considering the effects of social interactions:

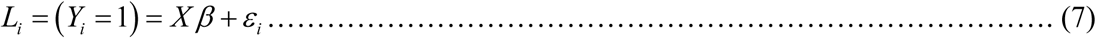

Where **X** represents the vector of explanatory variables (including the child’s age, gender, caregiver’s education, caregiver’s age, household size, household wealth, knowledge of ACT, perceptions of malaria treatment, and attitudes toward malaria treatment), **β** is the vector of parameters, and ε is the error term.

Secondly, we now estimate the binary logit regression model in equation (8) below which incorporates social interaction factors.

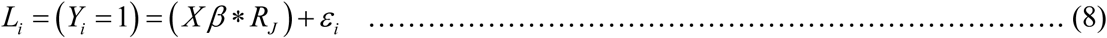

Where (**Xβ** * **R_J_**) includes interaction terms between the explanatory variables in equation (7) and the social interaction factors. **R** is a vector of the social interaction factors and **J** = (Christian or Muslim)

### Source of Data

This study uses cross-sectional household survey data from the 2020 Kenya Malaria Indicator Survey (KMIS). ^[17]^ The dataset consists of clusters of various sizes and populations, with each survey period showing an increase in the number of clusters included in KMIS. In rural areas, clusters are typically villages or groups of villages, while in urban settings, they are city blocks. The 2020 KMIS survey was nationally representative, covering a total of 30,252 weighted households. From this, a sample of 7,952 households was selected, and 6,771 women aged 15-49 years were interviewed, with a high response rate of 96% for women and 97% for households. This comprehensive dataset allows the study to explore not only the effectiveness of malaria treatments but also the influence of social interactions on treatment decisions within diverse environmental and socio-economic contexts.

### Ethical Consideration

The 2020 Kenya Malaria Indicator Survey (KMIS) followed strict ethical guidelines approved by the Kenyatta National Hospital/University of Nairobi Ethics Review Committee and ICF’s Institutional Review Board. Informed consent was obtained from participants after explaining the study’s objectives and potential risks. Participation was voluntary, with no incentives, and personal identifiers were removed to ensure confidentiality.

### Patient and Public Involvement

In the Kenya Malaria Indicator Survey (KMIS) 2020, patients or the public were not directly involved in the design, conduct, reporting, or dissemination plans of the research. KMIS studies are typically designed and implemented by national programs such as the Division of National Malaria Program (DNMP) in collaboration with organizations like the Kenya National Bureau of Statistics (KNBS) and international partners, including the U.S. President’s Malaria Initiative (PMI) and the World Health Organization (WHO). These organizations adhere to established methodologies for national health surveys, guided by international standards.

Although direct involvement of patients or the public is not explicitly mentioned, the surveys rely on household participation for data collection, which indirectly involves the public by capturing their experiences, practices, and outcomes regarding malaria prevention and treatment.

### Pre-estimation Tests

Several potential estimation issues can arise when fitting a binary logit model with multiple interaction terms using the Kenya Malaria Indicator Survey (KMIS) 2020 data set. This could include issues such as multicollinearity, model fit, specification errors, and the presence of influential observations could pose problems.

### Multicollinearity

The VIF is calculated for the model represented above in Equation (8) and results are presented in the Table 1. Results indicate that on average the model in the study exhibit low levels of multicollinearity with an average of 3.73 for usage of malaria medicines.

**Table 1:** VIF Uncentered Estimates with the Dependent Variable being the Uptake of ACT/other Drugs.

| **Explanatory Variables** | **VIF Uncentered** | **1/VIF** |
| --- | --- | --- |
| Child gender | 1.80 | 0.555787 |

| **Child Age in years** |  |  |
| --- | --- | --- |
| 1 | 2.32 | 0.431212 |
| 2 | 2.67 | 0.374205 |
| 3 | 2.65 | 0.377837 |
| 4 | 2.56 | 0.390768 |
| 5 | 2.25 | 0.443929 |
| Age of child caregiver | 17.77 | 0.056268 |
| **Education of child caregiver** |  |  |
| Primary | 5.04 | 0.198421 |
| Secondary | 4.54 | 0.220109 |
| Higher | 2.76 | 0.361799 |
| Household size | 6.98 | 0.143309 |
| **Household wealth indicator** |  |  |
| Poorer | 1.69 | 0.593393 |
| Middle-income | 2.32 | 0.431636 |
| Richer | 4.17 | 0.239986 |
| Richest | 4.01 | 0.249640 |
| Knowledge of ACT | 1.18 | 0.847418 |
| Perception of malaria treatment | 1.25 | 0.800873 |
| Attitude toward malaria treatment | 1.22 | 0.822905 |
| **Mean VIF** | 3.73 |  |

**Source: Author**

### Specification Errors

The model for the uptake of malaria medicines in the study is based also on a large sample size of 15,186 which generally helps ensure more reliable and robust results. The high values of LR chi2 of 2980.51 in the linktest as shown in Table 2 indicate that the model significantly improves the fit over the null models. The p-value is below 0.05, indicating that the overall models are statistically significant.

**Table 2:** Linktest Estimates to Check for Omitted Variable Bias and Correct Model Specification.

| **Dependent variable** | **Coefficient** | **p-values** |
| --- | --- | --- |
| **Uptake of ACT** |  |  |
| _hat | 1.1144 (0.05) | 0.000 |
| _hatsq | 0.0406(0.01) | 0.006 |
| Constant | 0.0301(0.04) | 0.396 |
| LR chi2(2) | 2980.51 |  |
| Prob>chi2 | 0.0000 |  |
| Pseudo R2 | 0.2041 |  |
| **Number of observations** | **15,186** |  |

**Source: Author**

## Empirical findings

### Summary Statistics

This section provides an overview of the descriptive statistics for the analytical sample derived from the 2020 Kenya Malaria Indicator Survey (KMIS) data. These statistics provide valuable insights into various aspects of malaria treatment, as well as individual and household characteristics. The data is analyzed across three levels: individual, residential (representing the reference area’s geographic and environmental context), and religious (capturing social interaction factors) as shown in Table 3.

**Table 3:**
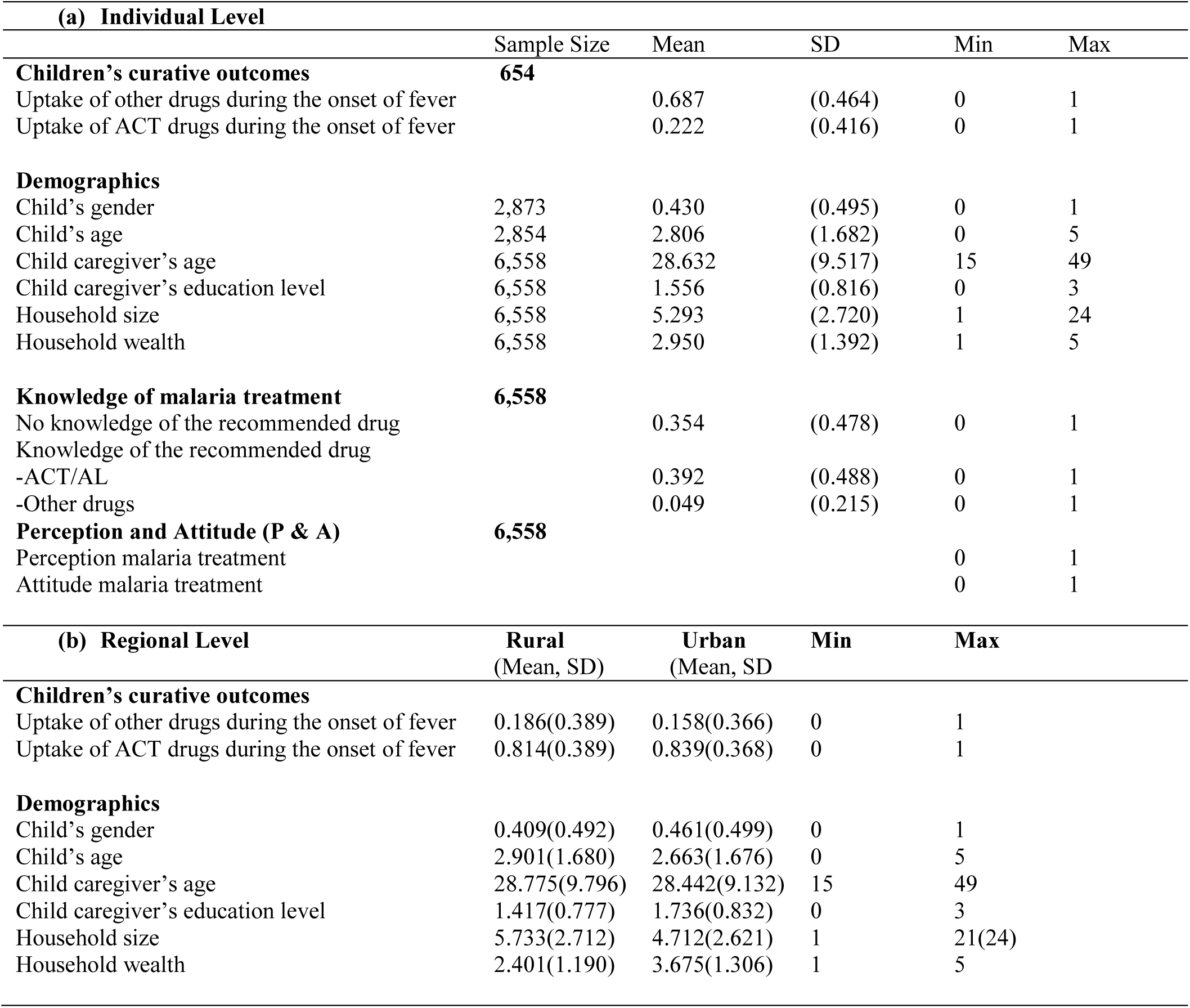
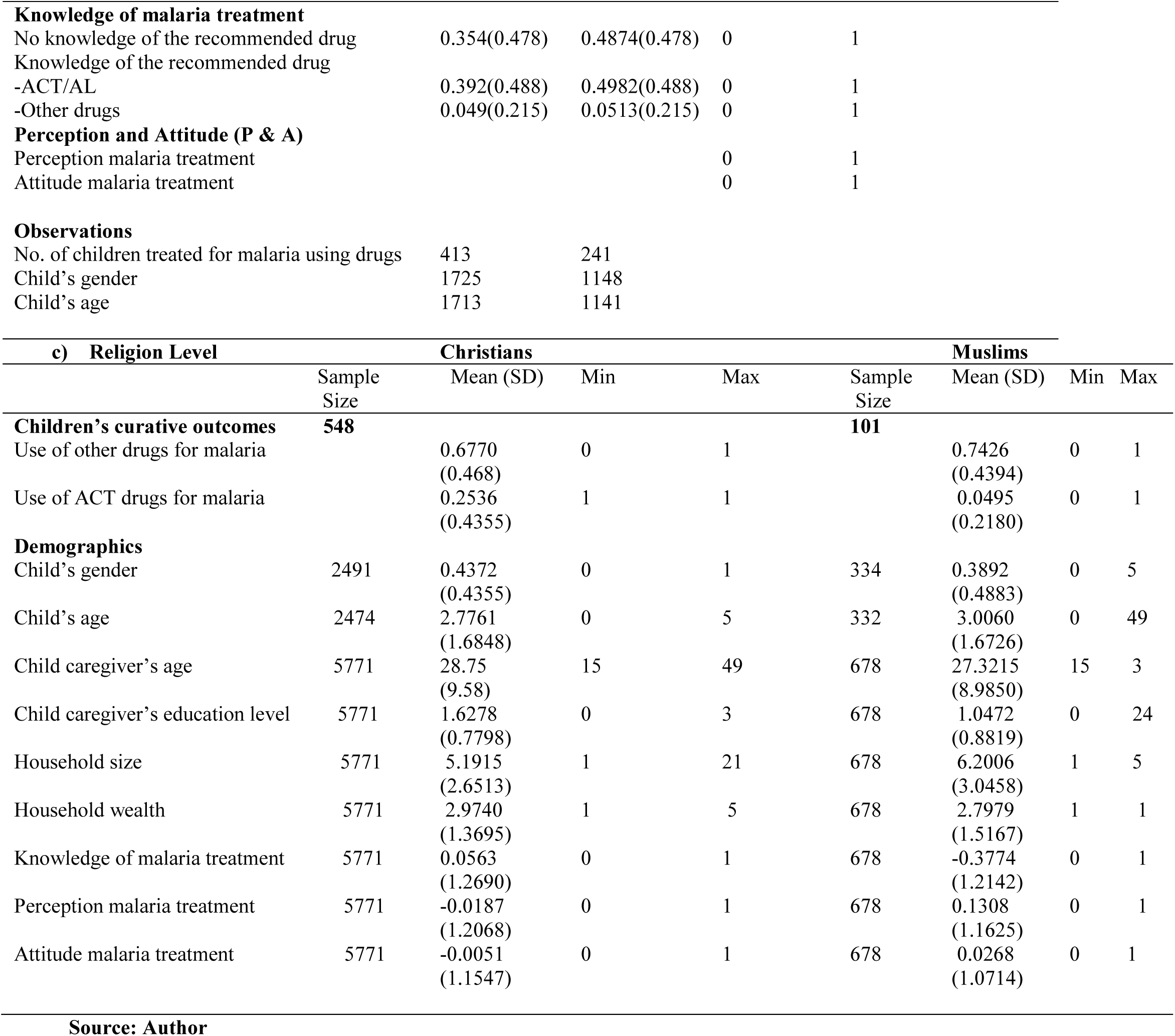
Descriptive Statistics of Variables used in the Estimations.

| **(a) Individual Level** |  |  |  |  |  |
| --- | --- | --- | --- | --- | --- |
|  | Sample Size | Mean | SD | Min | Max |
| **Children's curative outcomes** | **654** |  |  |  |  |
| Uptake of other drugs during the onset of fever |  | 0.687 | (0.464) | 0 | 1 |
| Uptake of ACT drugs during the onset of fever |  | 0.222 | (0.416) | 0 | 1 |
| **Demographics** |  |  |  |  |  |
| Child's gender | 2,873 | 0.430 | (0.495) | 0 | 1 |
| Child's age | 2,854 | 2.806 | (1.682) | 0 | 5 |
| Child caregiver's age | 6,558 | 28.632 | (9.517) | 15 | 49 |
| Child caregiver's education level | 6,558 | 1.556 | (0.816) | 0 | 3 |
| Household size | 6,558 | 5.293 | (2.720) | 1 | 24 |
| Household wealth | 6,558 | 2.950 | (1.392) | 1 | 5 |
| **Knowledge of malaria treatment** | **6,558** |  |  |  |  |
| No knowledge of the recommended drug |  | 0.354 | (0.478) | 0 | 1 |
| Knowledge of the recommended drug |  |  |  |  |  |
| -ACT/AL |  | 0.392 | (0.488) | 0 | 1 |
| -Other drugs |  | 0.049 | (0.215) | 0 | 1 |
| **Perception and Attitude (P & A)** | **6,558** |  |  |  |  |
| Perception malaria treatment |  |  |  | 0 | 1 |
| Attitude malaria treatment |  |  |  | 0 | 1 |
| **(b) Regional Level** |  |  |  |  |  |
|  | Rural<br>(Mean, SD) | Urban<br>(Mean, SD) | Min | Max |  |
| **Children's curative outcomes** |  |  |  |  |  |
| Uptake of other drugs during the onset of fever | 0.186(0.389) | 0.158(0.366) | 0 | 1 |  |
| Uptake of ACT drugs during the onset of fever | 0.814(0.389) | 0.839(0.368) | 0 | 1 |  |
| **Demographics** |  |  |  |  |  |
| Child's gender | 0.409(0.492) | 0.461(0.499) | 0 | 1 |  |
| Child's age | 2.901(1.680) | 2.663(1.676) | 0 | 5 |  |
| Child caregiver's age | 28.775(9.796) | 28.442(9.132) | 15 | 49 |  |
| Child caregiver's education level | 1.417(0.777) | 1.736(0.832) | 0 | 3 |  |
| Household size | 5.733(2.712) | 4.712(2.621) | 1 | 21(24) |  |
| Household wealth | 2.401(1.190) | 3.675(1.306) | 1 | 5 |  |

|  |  |  |  |  |
| --- | --- | --- | --- | --- |
| **Knowledge of malaria treatment** |  |  |  |  |
| No knowledge of the recommended drug | 0.354(0.478) | 0.4874(0.478) | 0 | 1 |
| Knowledge of the recommended drug |  |  |  |  |
| -ACT/AL | 0.392(0.488) | 0.4982(0.488) | 0 | 1 |
| -Other drugs | 0.049(0.215) | 0.0513(0.215) | 0 | 1 |
| **Perception and Attitude (P & A)** |  |  |  |  |
| Perception malaria treatment |  |  | 0 | 1 |
| Attitude malaria treatment |  |  | 0 | 1 |
| **Observations** |  |  |  |  |
| No. of children treated for malaria using drugs | 413 | 241 |  |  |
| Child's gender | 1725 | 1148 |  |  |
| Child's age | 1713 | 1141 |  |  |

| c) Religion Level |  | Christians |  |  | Muslims |  |  |  |
| --- | --- | --- | --- | --- | --- | --- | --- | --- |
|  | Sample Size | Mean (SD) | Min | Max | Sample Size | Mean (SD) | Min | Max |
| **Children's curative outcomes** | **548** |  |  |  | **101** |  |  |  |
| Use of other drugs for malaria |  | 0.6770<br>(0.468) | 0 | 1 |  | 0.7426<br>(0.4394) | 0 | 1 |
| Use of ACT drugs for malaria |  | 0.2536<br>(0.4355) | 1 | 1 |  | 0.0495<br>(0.2180) | 0 | 1 |
| **Demographics** |  |  |  |  |  |  |  |  |
| Child's gender | 2491 | 0.4372<br>(0.4355) | 0 | 1 | 334 | 0.3892<br>(0.4883) | 0 | 5 |
| Child's age | 2474 | 2.7761<br>(1.6848) | 0 | 5 | 332 | 3.0060<br>(1.6726) | 0 | 49 |
| Child caregiver's age | 5771 | 28.75<br>(9.58) | 15 | 49 | 678 | 27.3215<br>(8.9850) | 15 | 3 |
| Child caregiver's education level | 5771 | 1.6278<br>(0.7798) | 0 | 3 | 678 | 1.0472<br>(0.8819) | 0 | 24 |
| Household size | 5771 | 5.1915<br>(2.6513) | 1 | 21 | 678 | 6.2006<br>(3.0458) | 1 | 5 |
| Household wealth | 5771 | 2.9740<br>(1.3695) | 1 | 5 | 678 | 2.7979<br>(1.5167) | 1 | 1 |
| Knowledge of malaria treatment | 5771 | 0.0563<br>(1.2690) | 0 | 1 | 678 | -0.3774<br>(1.2142) | 0 | 1 |
| Perception malaria treatment | 5771 | -0.0187<br>(1.2068) | 0 | 1 | 678 | 0.1308<br>(1.1625) | 0 | 1 |
| Attitude malaria treatment | 5771 | -0.0051<br>(1.1547) | 0 | 1 | 678 | 0.0268<br>(1.0714) | 0 | 1 |

Source: Author

The sample includes 43.0% male children, with an average age of 2.8 years as indicated in section b. The mean age of child caregivers or pregnant women is 28.6 years, spanning from 15 to 49 years. On average, the education level of these caregivers is around 1.6 years. The average household size is about 5.3 members, with a range from 1 to 24 members. The average household wealth index is around 3, placing it within the middle-income group on a scale where 1 denotes the poorest households and 5 represents the wealthiest. On average, 35.4% of individuals did not know the recommended drug for malaria treatment, while 39.2% knew about artemisinin-based combination therapy/artemether-lumefantrine as the recommended treatment. Finally, only 4.9% of individuals were aware of other recommended drugs for malaria treatment.

Section b indicate that 18.6% of rural children received other drugs for malaria treatment, while this figure was 15.8% for urban children. Comparing demographic factors, 40.9% of the children under five years old in the survey are from rural areas, while 46.1% are from urban areas. The average age of children under five in rural areas is 2.9 years, while in urban areas it is 2.7 years. The mean age of child caregivers differs by 0.4 years between rural and urban areas, with a 0.3-year difference in education levels. The average household size is higher in rural areas, with 5.7 members, compared to 4.7 members in urban areas. The average wealth index is higher in urban areas, at 3.7, compared to 2.4 in rural areas. In rural areas, 35.4% of child caregivers lack knowledge of the recommended drugs for malaria treatment, while 39.2% are aware that ACT is the recommended drug. In urban areas, 47.8% of caregivers do not know the recommended drug, while 48.8% are aware that ACT is the recommended drug. Lastly, 4.9% of rural caregivers are unaware of other recommended drugs for malaria treatment, compared to 21.5% in urban areas.

Section c provides a summary of the variables concerning religious affiliations categorized as Christians and Muslims. The Christian variable includes the combined sum of Catholics and Protestants in the dataset. These analyses offer insights into the differences between Christians and Muslims regarding various factors related to malaria treatment and demographics. A greater proportion of children under five in Christian and Muslim communities (67.7% and 74.3% respectively) were given other drugs on the onset of fever while 25.4% and 5% respectively were given the recommended ACT drug on the onset of fever.

## Results

The findings on the uptake of artemisinin-based combination therapy (ACT) by children under five reveal significant variations based on rural versus urban residence and religious affiliation (Christianity and Islam) are shown in Tables 4.

**Table 4:**
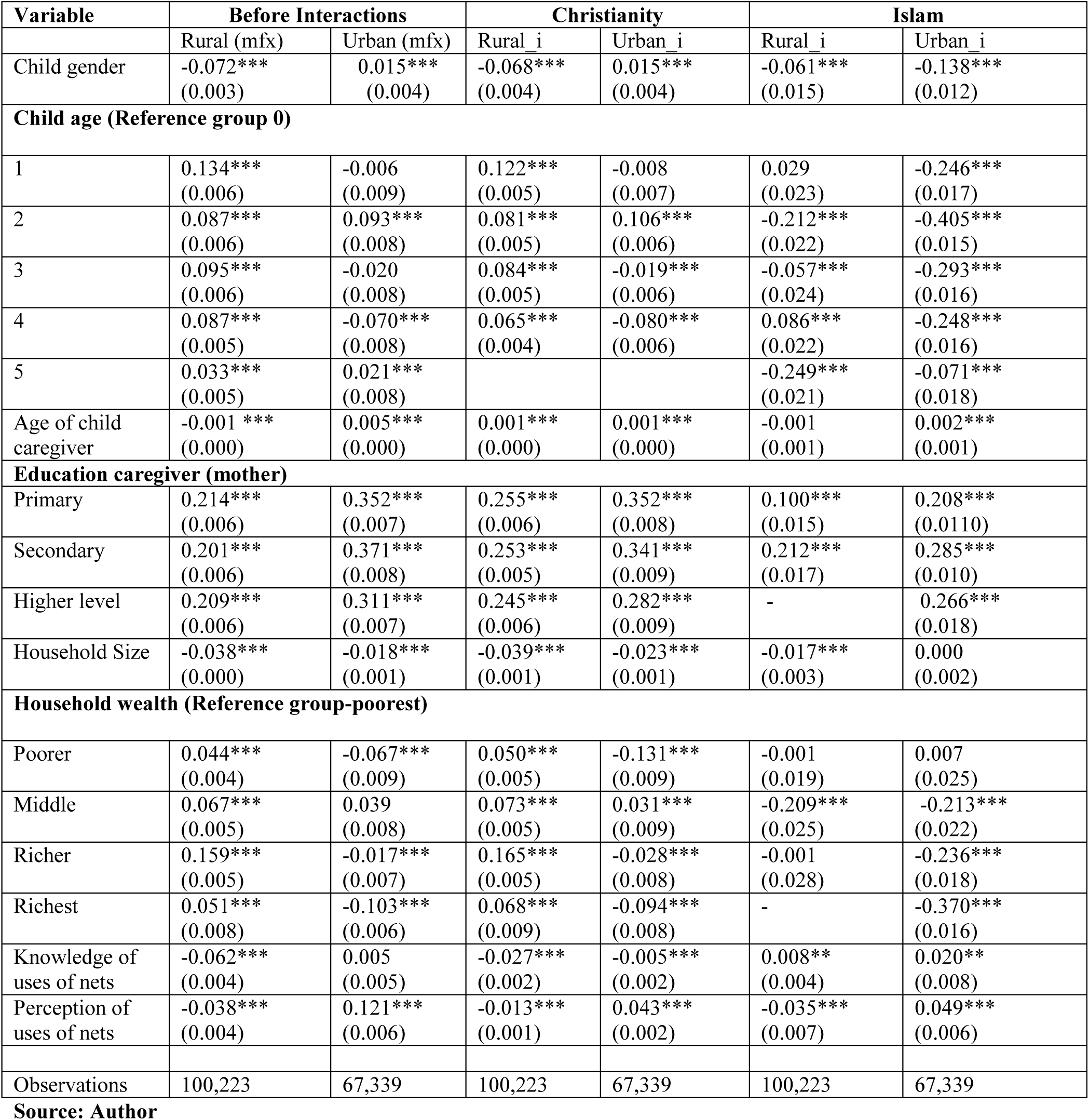
Logistic Regression Estimation results of the Uptake of ACT Drugs by Children Under 5 Years by Residence in Kenya, 2020.

| Variable | Before Interactions |  | Christianity |  | Islam |  |
| --- | --- | --- | --- | --- | --- | --- |
|  | Rural (mfx) | Urban (mfx) | Rural_i | Urban_i | Rural_i | Urban_i |
| Child gender | -0.072***<br>(0.003) | 0.015***<br>(0.004) | -0.068***<br>(0.004) | 0.015***<br>(0.004) | -0.061***<br>(0.015) | -0.138***<br>(0.012) |
| **Child age (Reference group 0)** |  |  |  |  |  |  |
| 1 | 0.134***<br>(0.006) | -0.006<br>(0.009) | 0.122***<br>(0.005) | -0.008<br>(0.007) | 0.029<br>(0.023) | -0.246***<br>(0.017) |
| 2 | 0.087***<br>(0.006) | 0.093***<br>(0.008) | 0.081***<br>(0.005) | 0.106***<br>(0.006) | -0.212***<br>(0.022) | -0.405***<br>(0.015) |
| 3 | 0.095***<br>(0.006) | -0.020<br>(0.008) | 0.084***<br>(0.005) | -0.019***<br>(0.006) | -0.057***<br>(0.024) | -0.293***<br>(0.016) |
| 4 | 0.087***<br>(0.005) | -0.070***<br>(0.008) | 0.065***<br>(0.004) | -0.080***<br>(0.006) | 0.086***<br>(0.022) | -0.248***<br>(0.016) |
| 5 | 0.033***<br>(0.005) | 0.021***<br>(0.008) |  |  | -0.249***<br>(0.021) | -0.071***<br>(0.018) |
| Age of child caregiver | -0.001***<br>(0.000) | 0.005***<br>(0.000) | 0.001***<br>(0.000) | 0.001***<br>(0.000) | -0.001<br>(0.001) | 0.002***<br>(0.001) |
| **Education caregiver (mother)** |  |  |  |  |  |  |
| Primary | 0.214***<br>(0.006) | 0.352***<br>(0.007) | 0.255***<br>(0.006) | 0.352***<br>(0.008) | 0.100***<br>(0.015) | 0.208***<br>(0.0110) |
| Secondary | 0.201***<br>(0.006) | 0.371***<br>(0.008) | 0.253***<br>(0.005) | 0.341***<br>(0.009) | 0.212***<br>(0.017) | 0.285***<br>(0.010) |
| Higher level | 0.209***<br>(0.006) | 0.311***<br>(0.007) | 0.245***<br>(0.006) | 0.282***<br>(0.009) | - | 0.266***<br>(0.018) |
| Household Size | -0.038***<br>(0.000) | -0.018***<br>(0.001) | -0.039***<br>(0.001) | -0.023***<br>(0.001) | -0.017***<br>(0.003) | 0.000<br>(0.002) |
| **Household wealth (Reference group-poorest)** |  |  |  |  |  |  |
| Poorer | 0.044***<br>(0.004) | -0.067***<br>(0.009) | 0.050***<br>(0.005) | -0.131***<br>(0.009) | -0.001<br>(0.019) | 0.007<br>(0.025) |
| Middle | 0.067***<br>(0.005) | 0.039<br>(0.008) | 0.073***<br>(0.005) | 0.031***<br>(0.009) | -0.209***<br>(0.025) | -0.213***<br>(0.022) |
| Richer | 0.159***<br>(0.005) | -0.017***<br>(0.007) | 0.165***<br>(0.005) | -0.028***<br>(0.008) | -0.001<br>(0.028) | -0.236***<br>(0.018) |
| Richest | 0.051***<br>(0.008) | -0.103***<br>(0.006) | 0.068***<br>(0.009) | -0.094***<br>(0.008) | - | -0.370***<br>(0.016) |
| Knowledge of uses of nets | -0.062***<br>(0.004) | 0.005<br>(0.005) | -0.027***<br>(0.002) | -0.005***<br>(0.002) | 0.008**<br>(0.004) | 0.020**<br>(0.008) |
| Perception of uses of nets | -0.038***<br>(0.004) | 0.121***<br>(0.006) | -0.013***<br>(0.001) | 0.043***<br>(0.002) | -0.035***<br>(0.007) | 0.049***<br>(0.006) |
| Observations | 100,223 | 67,339 | 100,223 | 67,339 | 100,223 | 67,339 |

**Source: Author**

### Urban vs. Rural Differences in ACT Uptake

The analysis identified notable disparities in ACT uptake between rural and urban areas. In rural regions, male children were less likely to receive ACT treatment (-0.072, p<0.001), whereas gender had a positive impact on treatment uptake in urban settings (+0.015, p<0.001). Age also played a significant role, with younger children (particularly those aged 1 to 2 years) in rural areas having a higher likelihood of receiving ACT (age 1: +0.134, p<0.001; age 2: +0.087, p< 0.001) compared to their urban counterparts. Additionally, household wealth influenced ACT usage differently across settings; wealthier households in rural areas were more inclined to use ACT (middle-income households: +0.067, p<0.001), while in urban areas, higher household wealth was associated with reduced ACT uptake (-0.017, p<0.001). These findings suggest that socio-economic and demographic factors differentially impact malaria treatment practices across rural and urban contexts

### Influence of Religious Affiliations

The analysis highlighted the significant impact of religious affiliations on ACT uptake. In rural Christian households, the education level of caregivers was a strong determinant, with those having secondary education being more likely to seek ACT treatment for their children (+0.253, p<0.001). Conversely, in predominantly Muslim rural areas, factors such as child age and household wealth had a negative influence on ACT uptake (e.g., age 2:-0.212, p<0.001; richest households:-0.370, p<0.001). This suggests that social norms and health-seeking behaviors differ between religious groups, influencing the prioritization of malaria treatment in these communities.

### Role of Caregiver Education and Household Characteristics

Caregiver education and household characteristics were found to significantly impact ACT uptake. Mothers with higher levels of education, particularly in rural areas, were more likely to seek ACT treatment for their children (e.g., primary education: +0.214, p<0.001; secondary education: +0.201, p<0.001). However, larger household sizes were associated with lower ACT adoption in both rural and urban settings (-0.038, p<0.001), suggesting that limited resources in bigger households may hinder prompt access to malaria treatment. These findings highlight the importance of targeting educational interventions for caregivers and addressing household resource constraints to improve malaria treatment uptake.

## Discussion

Child gender significantly influences ACT uptake, with boys in rural Christian households more likely to receive treatment, while the effect is negative in Islamic households. Child age also plays a role, differing by religion and location: in Christian households, younger children in urban areas are more likely to receive ACT, whereas older children benefit more in rural settings. Conversely, Islamic households show a decline in ACT use among older children, especially those around five years old. These patterns underline the complex interplay between social, demographic, and religious factors affecting malaria treatment access, particularly in rural areas.

Maternal age and education further shape ACT uptake across religious groups. Older mothers in Christian households show minimal positive effects on treatment, mainly in urban settings, while younger mothers in rural areas show a slight negative effect. In contrast, older mothers in rural Islamic households are significantly more likely to support ACT uptake. Education also impacts uptake; primary education boosts ACT use in rural Islamic households, while higher education levels in Christian households correlate with reduced uptake, highlighting the nuanced influence of education on healthcare decisions.

Caregiver perceptions and attitudes toward malaria treatment are crucial, particularly in rural households, where positive attitudes correlate with increased medication use. In urban areas, negative perceptions pose challenges, underscoring the importance of targeted health education to address barriers to care. These findings align with broader studies (e.g., Romay-Barja et al., 2015), ^[18]^ which emphasize the role of caregiver knowledge in shaping treatment behaviors and the need for tailored interventions to improve malaria treatment access.

In rural Islamic households, positive attitudes (+0.167) significantly enhance ACT uptake, while in urban areas, negative attitudes (-0.067) hinder treatment access, suggesting cultural or contextual barriers. Similar research in Ghana reinforces that caregiver attitudes and knowledge drive proactive healthcare-seeking behavior, highlighting the necessity of addressing context-specific barriers to enhance malaria control. ^[19]^

## Conclusion

Social interactions, including religious peer effects, play a crucial role in promoting the uptake of ACT for malaria treatment in Kenya. Addressing socio-cultural and economic barriers, particularly in rural areas, can lead to more effective malaria control strategies. Findings underscore the importance of addressing social, educational, and perceptual barriers to improve malaria treatment outcomes for children under five. Targeted interventions, particularly in urban Muslim settings, need to address negative attitudes and perceptions, while efforts in rural areas can capitalize on enhancing caregiver education and positive social interactions. Promoting awareness about ACT as the recommended treatment, alongside addressing the nuanced drivers of medication uptake, will be vital in achieving more equitable access to effective malaria care for all children in these contexts. Future research should explore longitudinal approaches to better understand the long-term impact of social networks on health behaviors

### Policy Implications

To enhance malaria management in children under five, ensuring consistent availability and accessibility of artemisinin-based combination therapies (ACTs) is crucial. This requires addressing supply chain disruptions and improving distribution networks to prevent stock-outs. Training healthcare providers on the timely and accurate administration of ACTs can also significantly impact treatment outcomes. Educational campaigns should focus on the benefits and correct usage of ACTs, with messaging tailored to align with local cultural and religious contexts to improve comprehension and adherence.

Research should investigate how social interactions, community leaders, and informal networks influence ACT uptake, as these factors can either facilitate or hinder adherence.

Addressing misinformation about antimalarial medications through targeted community engagement can help overcome barriers such as stigma and distrust in healthcare systems. Additionally, leveraging social media and digital platforms can support health behavior changes by developing technology-based interventions, such as mobile health applications and digital campaigns, to enhance adherence and facilitate access to ACTs.

## Declarations

## Authors’ Contributions

FNN, JKG, and GM were actively involved in the analysis and interpretation of the study’s findings. FNN drafted the initial version of the manuscript, while all authors contributed to revisions and approved the final version. FNN serves as the guarantor of the manuscript.

## Data Availability

All data produced are available online at https://statistics.knbs.or.ke/nada/index.php/catalog/111

## Acknowledgements

The authors express their sincere gratitude to the University of Nairobi and the Technical University of Kenya for their support and resources throughout this research journey. Special thanks go to the institutions responsible for the 2020 Kenya Malaria Indicator Survey (KMIS), the Ministry of Health, the Kenya National Bureau of Statistics (KNBS), the Division of National Malaria Programme (DNMP) and ther partners in malaria control for providing essential data that significantly contributed to this study. Your collective support has been instrumental in advancing this research on improving maternal health outcomes in malaria-prone regions.

## Ethical approval

The 2020 Kenya Malaria Indicator Survey (KMIS), was approved by the Kenyatta National Hospital Ethical Committee, Ministry of Health, Kenya.

## Data Availability

Kenya Ministry of Health (2020). Kenya Malaria Indicator Survey 2020 Final Report. Nairobi: National Malaria Control Programme (NMCP), Kenya National Bureau of Statistics (KNBS), and partners. Available at <u>KNBS</u> or <u>Statistics Kenya</u> https://statistics.knbs.or.ke/nada/index.php/catalog/111

